# High-intensity interval training combining rowing and cycling efficiently improves insulin sensitivity, body composition and VO_2_max in men with obesity and type 2 diabetes

**DOI:** 10.1101/2022.09.08.22279407

**Authors:** Maria Houborg Petersen, Martin Eisemann de Almeida, Emil Kleis Wentorf, Kurt Jensen, Niels Ørtenblad, Kurt Højlund

## Abstract

**Aim:** Non-weight-bearing high-intensity interval training (HIIT) involving several muscle groups may efficiently improve metabolic health in obesity and type 2 diabetes. In a non-randomized intervention study, we examined the effect of a HIIT-protocol, recruiting both lower and upper body muscles, on insulin sensitivity, measures of metabolic health and adherence in obesity and type 2 diabetes.

**Methods:** In 15 obese men with type 2 diabetes and age-matched obese (*n*=15) and lean (*n*=18) glucose-tolerant men, the effects of 8-weeks supervised HIIT combining rowing and cycling were examined by DXA-scan, exercise test and hyperinsulinemic-euglycemic clamp.

**Results:** At baseline, insulin-stimulated glucose disposal rate (GDR) was ∼40% reduced in the diabetic vs the non-diabetic groups (all *p*<0.01). In response to HIIT, insulin-stimulated GDR increased ∼30-40% in all groups (all *p*<0.01) explained by increased glucose storage. These changes were accompanied by ∼8-15% increases in VO_2_max, (all *p*<0.01), decreased fat mass and increased lean body mass in all groups (all *p*<0.05). There were no correlations between these training adaptations and no group-differences in these responses. HbA1c showed a clinically relevant decrease in men with type 2 diabetes (4±2 mmol/mol; *p*<0.05). Importantly, adherence was high (>95%) and no injuries were reported.

**Conclusions:** A novel HIIT-protocol recruiting lower and upper body muscles efficiently improves insulin sensitivity, VO_2_max and body composition with intact responses in men with obesity and type 2 diabetes. The high adherence and lack of injuries show that non-weight-bearing HIIT involving several muscle groups is a promising mode of exercise training in obesity and type 2 diabetes.

## INTRODUCTION

The global prevalence of type 2 diabetes is rapidly increasing with obesity and sedentary lifestyle as major risk factors. Type 2 diabetes and obesity are characterized by insulin resistance, which is associated with an increased risk of cardiovascular morbidity and mortality [1]. However, current treatment options for insulin resistance are very limited, except for weight loss and physical activity. At the metabolic level, insulin resistance in obesity and type 2 diabetes is characterized by reduced insulin-stimulated glucose uptake and glucose storage in skeletal muscle [2-4]. At the molecular level, insulin resistance in skeletal muscle is associated with reduced insulin signaling to glucose transport and glycogen synthesis, accumulation of lipid metabolites and abnormalities in mitochondrial oxidative metabolism [3-7].

Physical activity plays an important role in the prevention and management of type 2 diabetes [8, 9]. Long term studies (>8 weeks) of exercise training have shown improvements in insulin sensitivity and glycemic control, together with beneficial changes in cardiorespiratory fitness (VO_2_max), body composition, blood pressure, and lipid profile in patients with type 2 diabetes [4, 9-11]. The adaptations underlying the beneficial metabolic effects of exercise training, also in obesity and type 2 diabetes, include increased mitochondrial content and function and increased abundance of proteins involved in insulin-signaling to glucose uptake and glucose storage [4, 12-16]. In individuals with type 2 diabetes or obesity, the majority of studies have reported the effects of endurance training at moderate intensities on cycle ergometers or treadmills and resistance training, either separately or in combination [9]. Moreover, in most studies, patients with type 2 diabetes have been compared only with weight and age-matched controls [2-4, 12, 13, 16]. This precludes the opportunity to find attenuated responses in these groups compared to lean, healthy individuals, and hence the potential presence of exercise resistance [17, 18]. Importantly, the most appropriate mode of exercise training for sedentary middle-aged individuals with obesity and type 2 diabetes, when it comes to beneficial effects on metabolic health, adherence and risk of injuries, remains to be established [10].

Lack of time is a frequent mentioned barrier to do exercise on a regular basis, also among adults with type 2 diabetes [10]. This explains why high-intensity interval training (HIIT) has gained increased interest recently. HIIT is defined as “near maximal” efforts, and is generally performed at an intensity higher than 80% of maximal heart rate. Indeed, HIIT-protocols with shorter training durations have shown the same or better effects on measures of insulin sensitivity, glucose metabolism, VO_2_max, and body composition compared with endurance training not only in healthy, lean individuals [19, 20], but also in obesity and type 2 diabetes [21-23]. However, in most HIIT studies, insulin sensitivity was determined as fasting or oral glucose tolerance test (OGTT) derived surrogate markers and not by the gold standard hyperinsulinemic-euglycemic clamp.

Skeletal muscle accounts for approximately 40% of total body weight and is the predominant site of insulin-stimulated glucose uptake [3]. Interestingly, a recent study indicates that recruitment of both upper and lower body muscle groups could be necessary to take full advantage of the insulin-sensitizing effect of exercise training [24]. Thus, while several previous studies using the one-leg technique have shown that insulin sensitivity is increased in the exercised muscles and not the inactive muscles [15, 25], it was recently reported that insulin-mediated glucose uptake is actually decreased in the non-exercised muscle groups [24]. However, previous studies of HIIT have mainly focused on lower body activities such as walking, running and cycling [19-23]. These findings suggest that the combination of rowing and cycling, which are both non-weight-bearing modes of exercise with a low risk of injuries, could enhance the beneficial effects of HIIT on insulin sensitivity and other measures of metabolic health by recruiting several lower and upper body muscle groups.

In the present study, we investigated the effects of a novel HIIT-protocol combing rowing and cycling on insulin sensitivity determined by the gold-standard hyperinsulinemic-euglycemic clamp, VO_2_max, body composition and substrate metabolism as well as adherence in obese men with type 2 diabetes, and whether these effects were attenuated compared with glucose-tolerant obese and lean glucose-tolerant men.

## MATERIAL AND METHODS

### Study population

The study was performed at Steno Diabetes Center Odense (SDCO) and Department of Sport Science and Clinical Biomechanics at the University of Southern Denmark (SDU) in the period from January 2018 to December 2019. Middle-aged (40-65 years) obese (BMI 27-36) men with type 2 diabetes (n=15) and glucose-tolerant obese (BMI 27-36; n=15) and lean (BMI 20-25; n=18) men were included in the study (Table 1). The participants were all non-smokers and matched according to age, and the obese men with or without type 2 diabetes were further matched according to BMI. Medication details and eligibility criteria are given in supporting information methods. Informed consent was obtained from all participants before inclusion. The study was approved by the Regional Scientific Ethical Committees for Southern Denmark and performed in accordance with the Helsinki Declaration II. Four participants did not complete the study. One lean person didn’t start the training period because of new knee problems, and one lean and two diabetic men withdrew their consent to continue training due to lack of time. Their baseline characteristics are included in the analysis. In this non-randomized intervention study, the changes in insulin sensitivity in the three groups response to the HIIT was the primary outcome, while HIIT-induced changes in measures of body composition, VO_2_max and substrate oxidation as well as adherence were secondary outcomes.

**Table 1.**
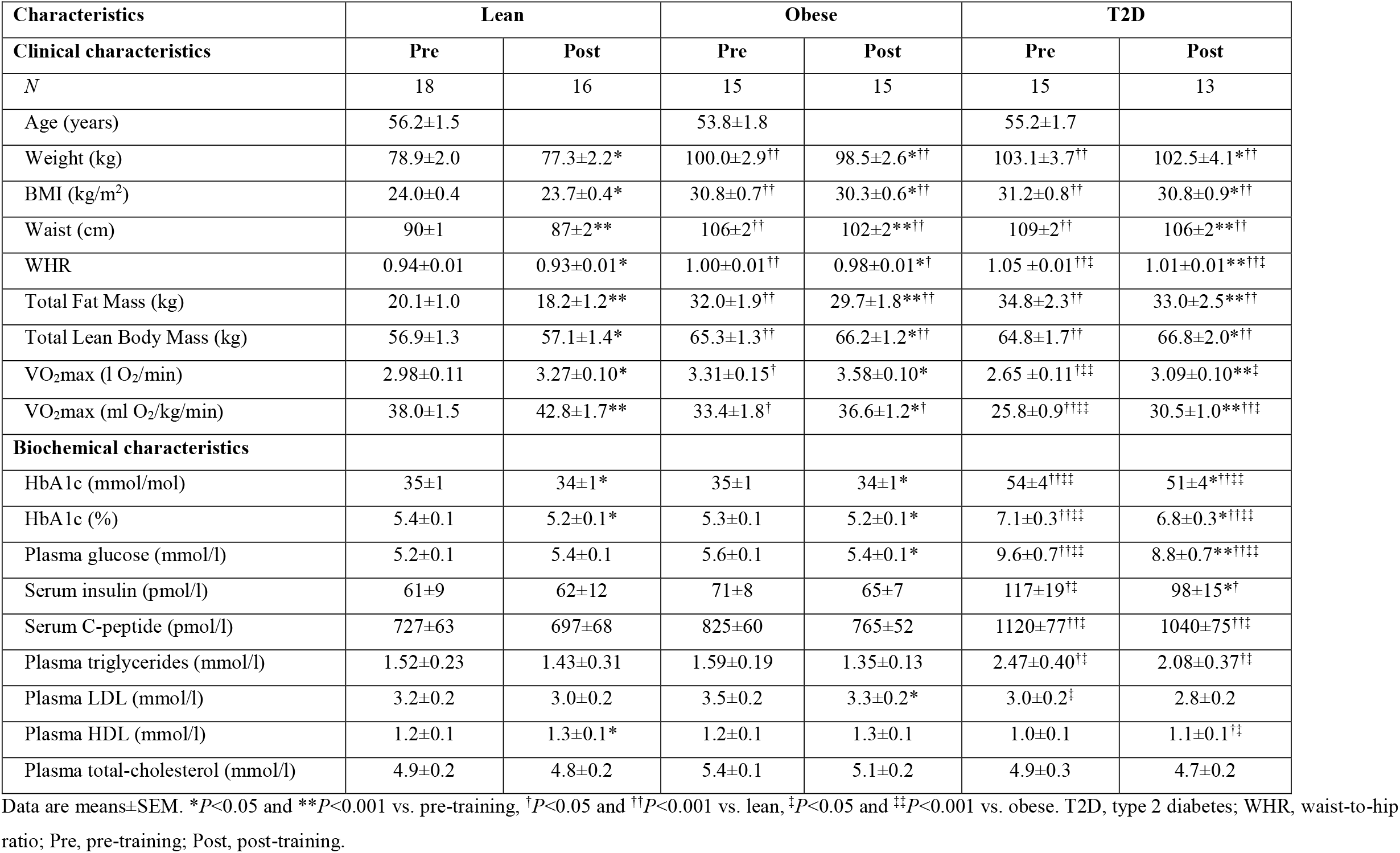
Clinical and biochemical characteristics, pre- and post-training.

### Study design

The study participants were examined on two days (Day 1 and 2) before separated by up to 2 weeks and on two days after (Day 3 and 4) an 8-week HIIT-protocol combining rowing and cycling (Fig. 1). On Day 1 and 3, total fat mass and lean body mass (LBM), maximal oxygen consumption (VO_2_max) and maximal cycling capacity (MCC) were determined (see supplementary material). On Day 2 and 4, biochemical characteristics and a hyperinsulinemic-euglycemic clamp combined with indirect calorimetry were performed to examine insulin sensitivity and substrate metabolism. Day 3 was performed 60 hours after the last afternoon HIIT-session, and Day 4 was performed 48 h after the VO_2_max test on Day 3. The participants were examined after an overnight fast and were instructed to refrain from strenuous physical activity for a period of 48-h and abstain from alcohol and caffeine for 24-h prior to experimental days. The participants were instructed not to change their dietary habits during the training intervention. In patients with type 2 diabetes, all medication was withdrawn one week prior to the clamp studies (Day 2 and 4), but was otherwise continued during the 8 weeks of HIIT.

**Fig. 1.**
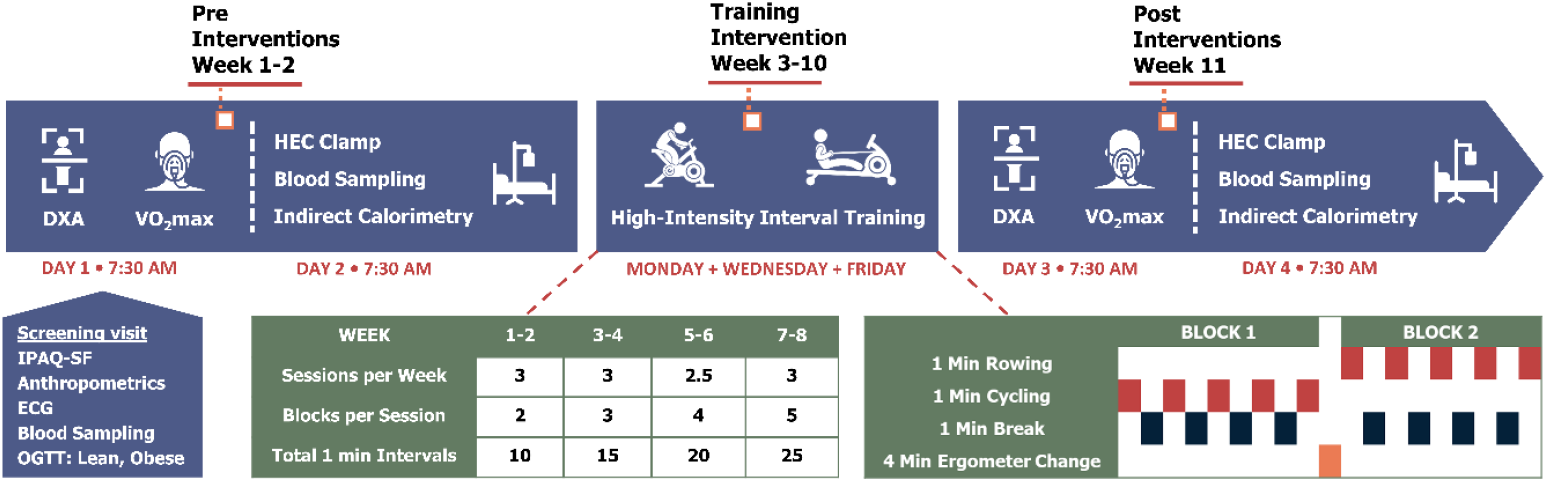
Study experimental design. HEC, hyperinsulinemic-euglycemic clamp; IPAQ-SF, International Physical Activity Questionnaire Short Form; OGTT, oral glucose tolerance test.

### Hyperinsulinemic-euglycemic clamp and biochemical analysis

Before (Day 2) and after (Day 4) the HIIT-protocol, the participants underwent a hyperinsulinemic-euglycemic clamp with tracer glucose consisting of a 2-h basal tracer equilibration period and a 3-h insulin-stimulated period using an insulin infusion rate of 40 mU/min/m^2^. For further details, see supplementary material. This was combined with indirect calorimetry to assess glucose infusion rate (GIR), tracer-determined glucose disposal rates (GDR) and hepatic glucose production (HGP), respiratory exchange ratio (RER), as well as rates of glucose (GOX) and lipid oxidation (LOX), and non-oxidative glucose metabolism (NOX) in the basal and insulin-stimulated steady-state periods as described (see also supplementary material) [2]. In patients with type 2 diabetes, plasma glucose was allowed to decline to 5.5 mmol/l before glucose infusion was initiated. Plasma glucose, triglycerides, and cholesterols, and serum insulin and C-peptide were measured as described (see supplementary material).

### HIIT protocol

In all participants, the effects of 8 weeks supervised HIIT combing rowing and cycling (3 sessions per week) were investigated. Participants in all three groups trained together in small groups of up to 10 participants. There was a continuous run-in and completion of participants. The majority of the training sessions (>95%) took place in the afternoon. Each training session started with a 10-min warm-up period on rowing (Concept II Model E, Vermont, US) or cycle ergometers (WattBike Pro/Trainer, Nottingham, UK), including row-specific technique training with increasing workload and heart rate. The HIIT-protocol consisted of training blocks of 5 × 1 min high-intensity intervals each interspersed by 1 min active or resting recovery. Between the training blocks, the participants had a 4-min break in which they shifted from rowing to cycling and vice versa. The number of blocks per session was gradually increased with an extra block added every second week, going from 2 blocks in week 1-2 to 5 blocks in week 7-8. The workload was set at 100-110% of MCC during cycling intervals, while the heart rate was kept similar at 86-88% of max during rowing and cycling. Participants were encouraged by social commitment, loud music, support from supervisors, and awareness of real-time relative heart rate due to monitoring (Polar H7, Polar team, Kempele, FI) throughout the sessions. Halfway through the HIIT-protocol, the workload was adjusted according to a midway VO_2_max test to ensure the relative workload was maintained throughout the training period.

### Deviations from protocol

Three men with type 2 diabetes and one lean man had their training period extended 1-1½ week due to illness during the training period. Two other lean and two obese men had their final DXA-scan and VO_2_max test (Day 3) 4-5 days after the last HIIT training session, and their final clamp (Day 4) 48-h after this.

### Statistics

All statistical analyses were performed by STATA/IC 16.1. The sample size were estimated to detect a 30% reduction in baseline insulin-stimulated GDR in men with T2D compared to lean, healthy men, and a 15% increase in insulin-stimulated GDR in men with T2D in response to HIIT. This gave a power of >80% when including 13 individuals in each group. Using mixed model linear regression, analyses for the HIIT-induced responses on outcome variables within each group were performed, as well as analyses comparing pre- and post-training data and the HIIT-induced responses between the groups. Data on self-reported levels of physical activity were compared between the groups by linear regression analyses (see supplementary material). Normality was checked using QQ-plots of residuals and for the mixed model also by QQ-plots of random effects. When residuals were not normally distributed, either Kruskal-Wallis test or binomial regression was performed. Pearson’s correlation coefficient was used for correlation analyses. Data are presented as means±SEM. Statistical significance was accepted at *p*<0.05.

## RESULTS

### Baseline characteristics

At baseline, HbA1c and fasting levels of plasma glucose, triglycerides and serum insulin and C-peptide were higher in men with type 2 diabetes than in both obese and lean men (Table 1), whereas body weight, BMI, waist, waist-hip-ratio (WHR), total fat mass and LBM were higher in both obese groups compared with the lean group. VO_2_max per kg body weight was lower in men with type 2 diabetes compared to both obese and lean men (all *p*<0.001), but also lower in obese than in lean men (*p*=0.02) (Table 1). However, the self-reported levels of physical activity were not different between the three groups (see supplementary material, Table S1).

Insulin-stimulated GDR and NOX were ∼40% and ∼49% lower, respectively (all *P*<0.01), whereas insulin-suppressed LOX was ∼75-90% higher (all *p*<0.05) in men with type 2 diabetes compared to both obese and lean men (Table 2). Insulin-stimulated RER was slightly lower in men with T2D compared with lean men, and the ability to increase RER in response to insulin (ΔRER) was reduced in men with T2D compared to both obese and lean men. No differences in GOX or HGP were observed between the groups.

**Table 2.**
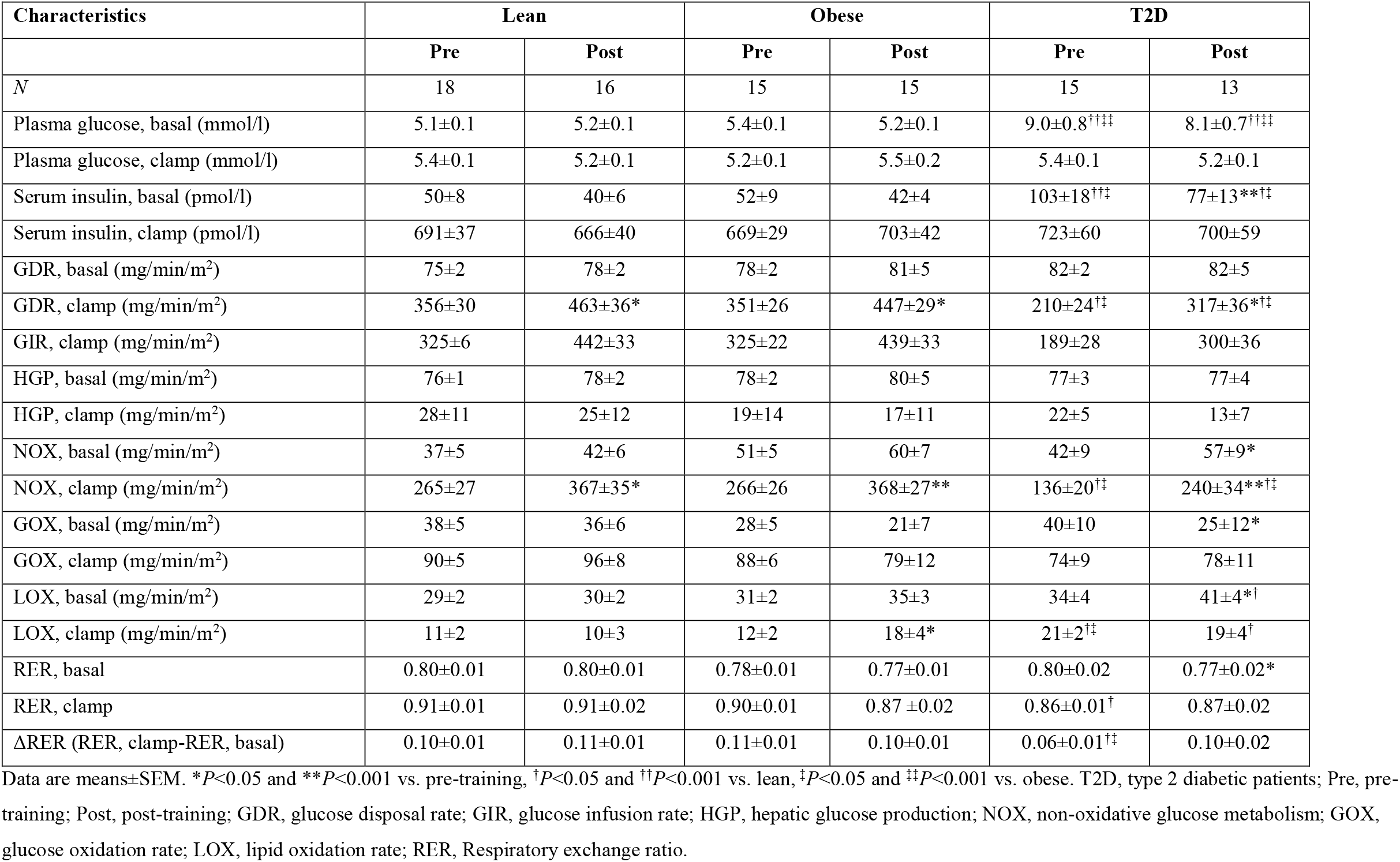
Metabolic characteristics during clamp, pre- and post-training.

### Compliance to the HIIT-protocol

The adherence was comparable between groups (all *P*>0.66), with an attendance rate above 95% in all three groups (see supplementary material, Table S2). There were no injuries reported during the 8-week HIIT. Although training volume increased from 2 to 5 blocks of 5 × 1-min intervals during the 8-week intervention, the participants in all three groups trained at a comparable intensity (% of max heart rate) throughout the training period, and managed to increase the workload in both cycling (2-3%) and rowing (7-10%) from week 1-4 to 5-8 (see supplementary material, Table S2).

### Effect of HIIT on VO_2_max and body composition

VO_2_max (l O_2_/min) increased by ∼15% in the diabetic group (*P*<0.001), ∼8% in the obese group (*p*=0.002) and ∼10% in the lean group (*p*=0.001) in response to HIIT (Table 1 and Fig 2b). There was considerable heterogeneity in the response of VO_2_max to HIIT in all groups (Fig 2f). However, only a few participants (*n*=5) did not increase VO_2_max. In all three groups, weight, BMI, waist and WHR were slightly reduced (all *p*<0.05) in response to HIIT (Table 1). The relatively small HIIT-induced reduction in weight was explained by a larger average decrease in total fat mass (1.6 to 2.3 kg; all *p*<0.05) than the increase in LBM (0.6 to 1.5 kg; all *p*<0.05) in each group (Table 1 and Fig. 2c-d). There were no differences in these HIIT-induced responses between the groups.

**Fig. 2.**
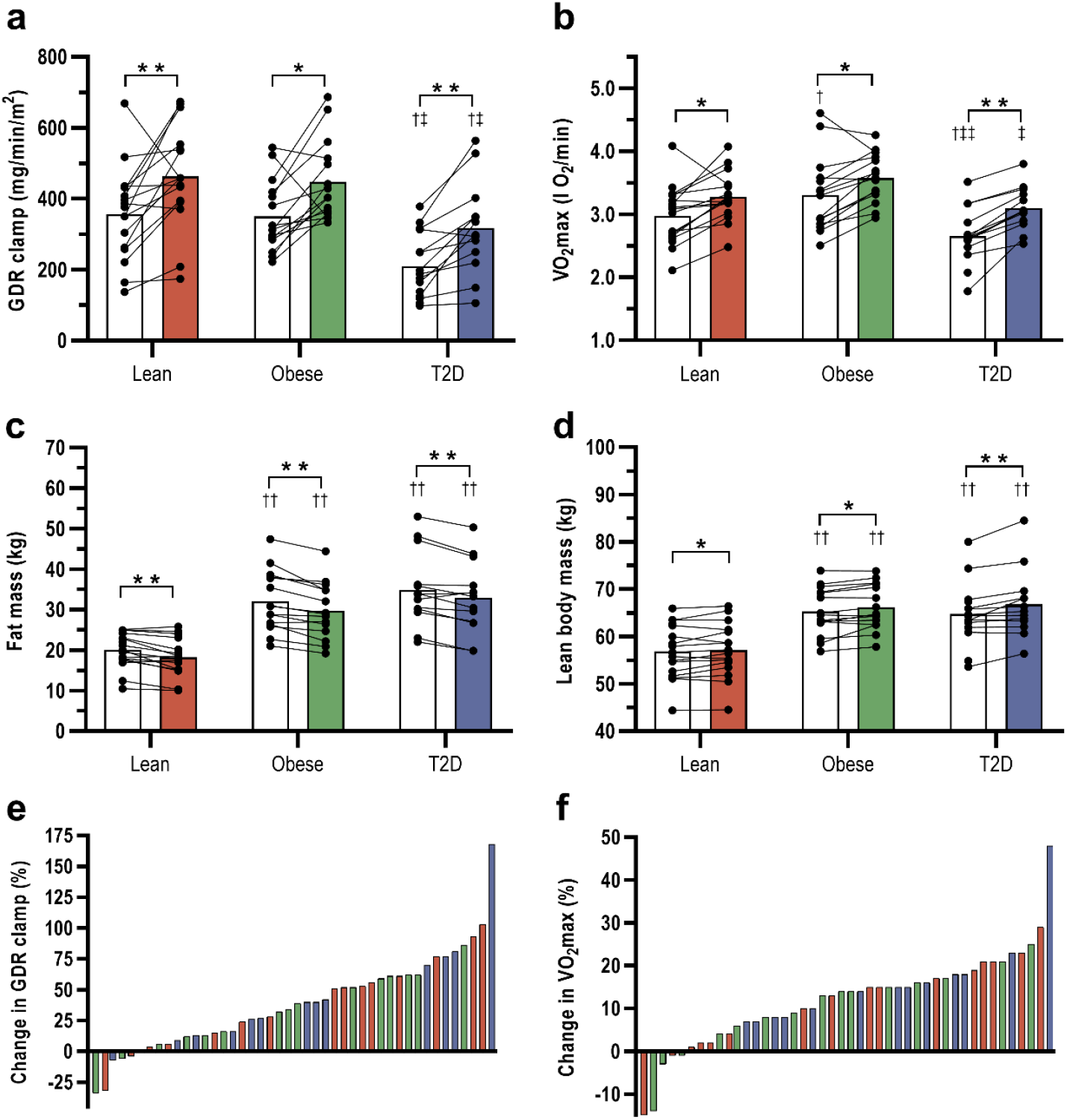
Effects of 8-weeks high-intensity interval training (HIIT) combining cycling and rowing on (*a*) insulin-stimulated glucose disposal rates (GDR clamp), (*b*) VO_2_max, (*c*) total fat mass, and (*d*) total lean body mass in patients with type 2 diabetes (T2D) and glucose tolerant obese and lean individuals. White bars show pre-training data and colored bars post-training data. (*e, f*) interindividual variability in the changes in GDR clamp and VO_2_max in response to HIIT in T2D (blue bars), obese (grey bars) and lean (red bars). Data are means±SEM. \**p*<0.05 and \*\**p*<0.001 vs. pre-training, †*p*<0.05 and ††*p*<0.001 vs. lean, ‡*p*<0.05 and ‡‡*p*<0.001 vs. obese.

### Glycemic control and lipid profile

HIIT induced a clinically relevant reduction in HbA1c (4±2 mmol/mol; (*p*=0.017) and fasting plasma glucose (1.0±0.4 mmol/l; *p*<0.001) in men with type 2 diabetes (Table 1). In obese and lean men, HbA1c and fasting plasma glucose were also slightly reduced in response to HIIT (Table 1), but the HIIT-induced reductions in HbA1c and fasting plasma glucose were larger in the diabetic versus the non-diabetic groups (all *p*<0.05). In response to HIIT, plasma HDL increased (*p*=0.002) in the lean group, and plasma LDL decreased (*p*=0.016) in the obese group, whereas plasma triglycerides and total cholesterol were unaffected by HIIT in all groups. There were no differences in these responses between the groups.

### Effect of HIIT on insulin sensitivity and substrate metabolism

In response to 8-week HIIT, insulin-stimulated GDR increased by 42% in men with type 2 diabetes (*p*<0.001), 27% in obese men (*p*=0.001) and 29% in lean men (*p*<0.001) (Table 2 and Fig 2a). There was a large interindividual variability in the response to HIIT in all three groups, but only a few participants (*n*=6) did not increase insulin-stimulated GDR (Fig. 2e). Post-training insulin-stimulated GDR was ∼30% lower in men with type 2 diabetes compared to both obese and lean men (*p*<0.05). However, post-training insulin-stimulated GDR in the diabetic group was not different from pre-training insulin-stimulated GDR in either the obese (*p*=0.400) or the lean group (*p*=0.483). The magnitude of the HIIT-induced increases in insulin-stimulated GDR was entirely explained by accompanying increases in insulin-stimulated NOX in all three groups (all *p*<0.001) as opposed to no HIIT-induced changes in insulin-stimulated GOX (Table 2). While insulin-suppressed LOX was unaffected by HIIT in men with type 2 diabetes and lean men, it increased in obese men. However, post-training insulin-suppressed LOX was only higher in men with type 2 diabetes compared with lean men (Table 2). Basal RER was slightly reduced in men with T2D in response to HIIT, and the attenuated ΔRER observed at baseline compared to the other groups were not present after HIIT. HIIT did not cause changes in HGP in any of the groups. There were no differences in any of the above HIIT-induced responses between the groups.

### Correlation analyses

To test whether changes in the different exercise response variables were correlated, we examined the relationship between the HIIT-induced changes in VO_2_max, insulin sensitivity, body composition and HbA1c. The HIIT-induced improvement in VO_2_max did not correlate with the increase in insulin-stimulated GDR either in the total cohort (*r*=-0.14, *p*=0.36) or any of the groups. Moreover, no correlations were found between the HIIT-induced increases in neither VO_2_max nor insulin-stimulated GDR and the changes in HbA1c, BMI, fat mass or LBM in the total cohort. However, the HIIT-induced increase in insulin-stimulated GDR correlated with the reduction in BMI (*r*=-0.61, *p*=0.027) and tended to correlate with the decrease in HbA1c (*r*=-0.51, *p*=0.077) in the diabetic group, but not in the lean and obese groups.

## DISCUSSION

In the present study, we examined the effects of a novel HIIT-protocol combining rowing and cycling in men with type 2 diabetes compared with glucose-tolerant obese and lean men. Our main finding is that this HIIT-protocol markedly improved insulin sensitivity with intact responses in obese men with and without type 2 diabetes. This was accompanied by, but did not correlate with, equally improved VO_2_max and body composition in all groups. Moreover, we found a clinically relevant decrease in HbA1c in patients with type 2 diabetes. Importantly, we observed a high adherence rate in each group and no injuries were reported. These results provide evidence that a supervised, non-weight-bearing HIIT-protocol involving a large muscle mass and several muscle groups is a promising mode of exercise training in sedentary individuals with obesity or type 2 diabetes.

We and others have previously reported increases in insulin-stimulated GDR by ∼10-20% in obese individuals with and without type 2 diabetes in response to 8-10 weeks supervised endurance training on cycle ergometers or treadmill [4, 26]. However, in young, sedentary lean men a similar 10-week supervised endurance training protocol increased insulin sensitivity by ∼30% [27], suggesting a lower response to exercise in obesity and type 2 diabetes as reported previously [18]. Here, we show that 8-weeks supervised HIIT combining rowing and cycling improved insulin sensitivity by ∼30-40% not only in lean glucose-tolerant men, but also in obesity and type 2 diabetes with the same magnitude. Thus, our HIIT-protocol increased insulin sensitivity ∼2-fold more than observed after endurance training in individuals with type 2 diabetes or obesity [4, 26], and, importantly, there was no evidence of exercise resistance. Interestingly, after completing the HIIT-protocol, the men with type 2 diabetes achieved a level of insulin sensitivity similar to obese and lean men at baseline.

Recent studies have compared the effect of HIIT with endurance training on insulin sensitivity in patients with type 2 diabetes using surrogate markers of insulin sensitivity such as HOMA-IR [23, 28]. In one study, 11-weeks of supervised HIIT on cycle ergometers reduced HOMA-IR by ∼20-25%, whereas no change was observed after endurance training [23]. In contrast, another study reported similar decreases in HOMA-IR by ∼20% after 12-weeks supervised HIIT and endurance training on treadmills [28]. Thus, it appears that HIIT has at least the same effect on HOMA-IR as endurance training in individuals with type 2 diabetes. However, only a few studies have investigated the effect of HIIT on insulin sensitivity using the hyperinsulinemic-euglycemic clamp. In young and older individuals, 6-12 weeks of supervised HIIT on cycle ergometers or treadmills increased insulin-stimulated glucose uptake by ∼11-20% [19, 20]. Taken together, these previous reports suggest that the 30-40% increase in insulin sensitivity observed in our study may not solely be ascribed to the choice of HIIT versus endurance training. Based on studies showing that insulin sensitivity is only increased in exercised muscles [15, 25], and perhaps even decreased in non-exercised muscle groups [24], we designed our HIIT-protocol to activate both lower and upper body muscle groups. Indeed, our results lend support to the hypothesis that recruitment of several muscle groups is necessary to take full advantage of the insulin-sensitizing effect of exercise training, and this may to a large extent explain the marked increase in insulin sensitivity in response to our HIIT-protocol. However, we cannot exclude a contribution from other differences in the design, e.g. afternoon training sessions [29], and longer durations of the HIIT-sessions during the last weeks compared with other HIIT-protocols performed at similar intensities [19-21].

Cardiorespiratory fitness, measured as VO_2_max, is considered a key response variable of exercise training on cardiovascular and metabolic health [30, 31]. Recent studies and meta-analyses have suggested a higher increase in VO_2_max in response to HIIT compared with endurance training in patients with type 2 diabetes [22, 23, 28, 32]. In the present study, our HIIT-protocol increased VO_2_max equally in all three sedentary groups with a magnitude (8-15%) comparable to that observed in several studies of both HIIT and endurance training [13, 19, 20, 33, 34]. Importantly, our findings demonstrate an intact HIIT-induced increase in VO_2_max in men with type 2 diabetes or obesity as compared to lean, sedentary individuals. However, in contrast to insulin sensitivity, we do not find a superior effect of HIIT or an additional effect of recruiting lower and upper body muscle groups on VO_2_max compared to previous endurance training studies (see above). This indicates that the metabolic response to this novel HIIT protocol goes beyond the effect on the cardiorespiratory fitness, which may be advantageous in the prevention and management of type 2 diabetes [30].

There is strong evidence that exercise training leads to variable responses or even a lack of response (non-response) in both healthy and diseased individuals [30, 31]. In line, we demonstrate a high interindividual variability in the VO_2_max response to HIIT in each group, ranging from a 15% decrease to a 48% increase. Interestingly, the HIIT-induced changes in insulin-stimulated GDR and body composition showed a similar high degree of heterogeneity suggesting an association between these responses in the study cohort. However, except for an association between the increase in insulin sensitivity and the reduction in BMI in patients with type 2 diabetes, we found no correlations between the HIIT-induced changes in VO_2_max, insulin sensitivity or body composition either in the total cohort or in the individual groups. These findings are consistent with the current view that an adaptation to exercise training in one outcome does not necessarily equal responses in other outcomes [30], and emphasize that the evaluation of an exercise training program should not rely on a single exercise response variable and herein VO_2_max.

Body composition plays an important role in cardiovascular and metabolic health. However, several studies of patients with type 2 diabetes have been unable to demonstrate an effect of endurance training on weight, BMI and body composition [4, 16, 18, 26]. Moreover, many studies comparing the effect of HIIT versus endurance training in patients with type 2 diabetes could not demonstrate superior effects of HIIT on weight, WHR, or body composition [23, 28, 33], although one study and recent meta-analyses indicate a better effect of HIIT on BMI and WHR [22, 32, 35]. A recent study of 8-weeks HIIT on cycle ergometers demonstrated reduced BMI and abdominal fat mass, but also an undesirable reduction in LBM in patients with type 2 diabetes [34]. In the present study, we demonstrate clinically relevant improvements of both total fat mass and LBM in response to our HIIT-protocol in men with type 2 diabetes and glucose-tolerant obese and lean men instructed to continue their habitual food intake. These changes were accompanied by smaller decreases in weight, BMI and WHR. Compared with the previous reports mentioned above, it appears likely that the recruitment of upper and lower body muscle groups rather than HIIT *per se* contributed to these clear improvements in body composition. As noted above, the changes in total fat mass and LBM did not correlate with the marked increase in insulin sensitivity suggesting other factors to be involved, e.g. intrinsic adaptations in skeletal muscle and adipose tissue function, which are not necessarily reflected by changes in muscle or fat mass.

A major challenge in the prevention and management of type 2 diabetes is a low engagement in physical activity [10, 36]. Although a high adherence to exercise training is critical to obtain the desired effects on metabolic health, it is often not reported or highly variable [36]. In our study, we observed a very high adherence to both targeted training intensity and training sessions in all three groups and no injuries were reported. The high degree of adherence is in part explained by the supervised training and lack of injuries [36], but also by fulfillment of reported motivators for exercise including enjoyment during exercise, training in small groups, achieving weight control, and use of heart rate monitors [37]. The lack of injuries could be explained by the non-weight-bearing mode of exercise, time for recovery between training sessions and a progressive training program. Supervised training of the type studied here is of course more costly than home-based training, but is most likely necessary to achieve a high adherence and thereby maintain the beneficial effects of exercise training on metabolic health.

The strengths of this study include 1) the inclusion of a lean, healthy group to rule out attenuated responses due to obesity or type 2 diabetes, 2) the application of a HIIT-protocol recruiting several lower and upper body muscle groups to maximize the insulin-sensitizing effect, 3) the supervised, group-based training sessions and the non-weight-bearing approach to ensure adherence and lack of injuries, and 4) conduction of the majority of the training sessions in the afternoon, which may enhance the metabolic benefits [29, 38]. However, this study also has limitations. First, we did not register the actual food intake of the participants and, therefore, cannot exclude that potential changes in their food intake influenced the results. Second, the duration of our HIIT-protocol may not capture the long-term effects of this training intervention. Third, the design of our study does not allow us to conclude whether the marked increase in insulin sensitivity was due to the HIIT-protocol, the recruitment of lower and upper body muscle groups or both. Finally, none of the men with type 2 diabetes included were on insulin treatment, and, our study only included men, which limits the generalizability of the results to women and type 2 diabetes patients with a higher degree of beta-cell failure.

### CONCLUSIONS

Our results demonstrate that a novel HIIT-protocol combining rowing and cycling efficiently improves insulin sensitivity, VO_2_max, and body composition with preserved responses in sedentary men with obesity and type 2 diabetes. The supervised training and non-weight-bearing mode of exercise ensured a high adherence rate and absence of injuries in all groups. In addition, the men with type 2 diabetes experienced a clinically relevant decrease in HbA1c. Our results also demonstrate that a supervised HIIT-protocol engaging lower and upper body muscle groups is a well-tolerated and promising mode of exercise training in sedentary individuals with obesity or type 2 diabetes. However, longer-term studies in larger cohorts including both men and women are needed to fully evaluate the metabolic and cardiovascular benefits of this HIIT protocol.

## Data Availability

All data produced in the present study are available upon reasonable request to the authors

## Abbreviations

FFA: Free fatty acid
GDR: Glucose disposal rate
GIR: Glucose infusion rate
GOX: Glucose oxidation
HGP: Hepatic glucose production
HIIT: High-intensity interval training
LBM: Lean body mass
LOX: Lipid oxidation
MCC: Maximal cycling capacity
NOX: Non-oxidative glucose metabolism
IPAQ: International physical activity questionnaire
RER: Respiratory exchange ratio
VO_2_: max Maximal oxygen consumption
W: Watt

## Acknowledgements

We would like to thank L. Hansen and C. B. Olsen at the Steno Diabetes Center Odense, Odense University Hospital for their skilled technical assistance. We thank J. V. Stidsen at the Steno Diabetes Center Odense, Odense University Hospital for analytical and statistical support. The Odense Patient data Explorative Network (OPEN) is acknowledged for statistical support and access to data storage in REDCap. Fig 1 has been designed using “Rowing Machine” and “Gym Cycling Machine” icons by Gan Khoon Lay from the Noun Project and resources from Flaticon.com.

## Authors contributions

M.H.P, K.J, N.Ø and K.H contributed to the conception and design of the study. M.H.P recruited the eligible participants and conducted the metabolic studies. M.E.A and E.K.W performed the supervised training, VO_2_max tests and DXA scanning. M.H.P, M.E.A, E.K.W, N.Ø and K.H analysed and interpreted data, and M.H.P and K.H wrote the manuscript. All authors have revised the manuscript critically for important intellectual content and given final approval of the version to be published. K.H and N.Ø are guarantors of this work, and as such had full access to all the data in the study and takes full responsibility for the integrity of the data and the accuracy of the data analysis.

