## Supplemental material for "High-intensity interval training combining rowing and cycling efficiently improves insulin sensitivity, body composition and VO_2_max in men with obesity and type 2 diabetes"

### SUPPLEMENTARY MATERIAL

### METHODS

#### Study population

Fifteen obese men with type 2 diabetes (duration 4.2±0.7 years) were carefully matched to 18 healthy, lean and 15 non-diabetic, obese men to participate in the study (Table 1). The sample sizes were estimated to detect a significant difference ($p<0.05$) in baseline insulin-stimulated GDR between patients with T2D and weight-matched controls, as well as a significant increase in insulin-stimulated GDR in response to exercise training in patients with T2D. This gave a power of >80%, when including at 13 subjects in each group. The level of physical activity for all participants had to be low or moderate (max 2 hours of moderate exercise weekly) and was evaluated by the short form of the International Physical Activity Questionnaire (IPAQ-SF) (see Table S1). Patients with type 2 diabetes were treated with metformin ($n$=14), DPP4-inhibitors ($n$=4) or sulphonylureas ($n$=1), either as monotherapy ($n$=11) or using two drugs ($n$=4). Patients with type 2 diabetes were all GAD65-antibody negative and had no signs of micro- or macrovascular complications, except for mild retinopathy ($n$=1), and most were treated with lipid lowering ($n$=11) and antihypertensive ($n$=12) medication. The lean and obese controls had normal glucose tolerance as evaluated by a 75 g OGTT and no family history of diabetes, and were not taking medication. All participants had normal results on blood screening tests for renal, hepatic and hematological function, and normal resting ECG. Informed consent was obtained from all participants before inclusion. The study was approved by the Regional Scientific Ethical Committees for Southern Denmark and performed in accordance with the Helsinki Declaration II. Four participants did not complete the study. Thus, one lean person didn’t start the training period because of new knee problems, and one lean and two diabetic men withdrew their consent to continue training due to lack of time. Their baseline characteristics are included in the analysis.

**Study design**

The study participants were examined on two days (Day 1 and 2) before (up to 2 weeks) and after (Day 3 and 4) a 8-week HIIT-protocol combining rowing and cycling (Fig. 1). On Day 1 and 3, total fat mass and lean body mass (LBM), maximal oxygen consumption ($VO_2$max) and maximal cycling capacity (MCC) were determined (See below). On Day 2 and 4, biochemical characteristics and a hyperinsulinemic-euglycemic clamp combined with indirect calorimetry were performed to examine insulin sensitivity and substrate metabolism. Day 3 was performed 3 days after the last HIIT-session, and Day 4 was performed 48 h after the $VO_2$max test on Day 3. The participants were examined after an overnight fast and were instructed to refrain from strenuous physical activity for a period of 48-h and abstain from alcohol, caffeine and nicotine 24-h prior to experimental days. The participants were instructed not to change their dietary habits during the training intervention. In patients with type 2 diabetes, all medication was withdrawn one week prior to the clamp studies (Day 2 and 4), but was continued during the 8 weeks of HIIT.

**Body composition and $VO_2$max**

At examination Day 1 and 3, the participants were served a standardized mixed breakfast meal (macronutrient composition; 64% carbohydrate, 22% fat and 14% protein) adjusted according to body weight. Total fat mass and lean body mass were assessed by dual-energy X-ray absorptiometry (DXA) scans (Prodigy Advance, GE Healthcare Lunar, Wisconsin, US). Approximately 1-2 h after consuming the mixed meal, $VO_2$max was determined by an incremental exercise test on a cycle ergometer (SRM Ergometer System, Jülich, DE) combined with a mixed chamber cardiopulmonary exercise analyzer (Oxigraf, Model O2CPX, California, US). After a warm up period, the workload was increased 20 watt (W) every 1 min until complete exhaustion was reached. Two out of following three criteria had to be achieved to ensure correct determination of $VO_2$max: 1) $VO_2$max reached a plateau despite further increases in workload, 2) blood-lactate, measured 2 min after ended test, was above 8.0 mmol/l and 3) the respiratory exchange ratio (RER) was greater than 1.10. Maximal cycling capacity (MCC) (=maximal power output) was calculated as the power output (W) before the last increase in workload, plus 20 W times the percentages of the 1 min cycled during the last increment [1].

**Hyperinsulinemic-euglycemic clamp and biochemical analysis**

Before (Day 2) and after (Day 4) the HIIT-protocol, the participants underwent a hyperinsulinemic-euglycemic clamp after an overnight fast as described previously [2-4]. The participants were instructed to eat a meal of similar size and macronutrient content in the evenings before the two clamps. In brief, the clamp consisted of a 2-h basal tracer equilibration period and a 3-h insulin-stimulated period using an insulin infusion rate of 40 mU/min/$m^2$. A primed primed-constant [3-3H]-glucose infusion was used throughout the clamp, and [3-3H]-glucose was added to the glucose infusates to maintain plasma specific activity constant at baseline levels during the 3-h insulin-stimulated clamp period as described [5]. Blood samples were drawn at baseline and every 10-20 min throughout the clamp. In patients with type 2 diabetes, plasma glucose was allowed to decline to 5.5 mmol/l during the insulin-stimulated period before glucose infusion was initiated. Total glucose disposal rates (GDR) and hepatic glucose production (HGP) were calculated using Steele´s non-steady-state equations during the final 40 min of the basal and insulin-stimulated steady-state periods [5]. The distribution volume of glucose was taken as 200 ml/kg body weight and pool fraction as 0.65. The clamp studies were combined with indirect calorimetry using a ventilated hood system (Oxycon Pro, Erich Jaeger GmbH, Hoechberg, Germany) to determine the respiratory exchange ratio (RER) and rates of glucose (GOX) and lipid oxidation (LOX) during the 30-min basal and insulin-stimulated steady-state periods as described [6]. Protein oxidation rates were estimated from urinary nitrogen excretion and corrected for changes in the pool size as described [7]. Rates of non-oxidative glucose metabolism (NOX) were calculated as the difference between GDR and GOX.

Plasma glucose was analyzed on Radiometer ABL800 FLEX Blood Gas Analyzer (Radiometer Medical ApS, Bronshoj, Denmark). Serum insulin and C-peptide levels were analyzed on Cobas e411 (Roche Diagnostics International Ltd., California). Plasma total cholesterol, HDL-cholesterol and -triglycerides were analyzed on heparinized plasma with absorption photometry on Cobas 8000 (Roche Diagnostics International Ltd., California).

### TABLES

#### Table S1 – Baseline physical activity and adherence to HIIT

| | **Lean** | **Obese** | **T2D** |
|---|---|---|---|
| *n* | 16 | 15 | 13 |
| **Adherence – week 1-8** | | | |
| HIIT sessions completed (%) | 97±1 | 98±1 | 95±1 |
| **IPAQ-SF baseline (time spent, h/week)** | | | |
| Vigorous physical activity | 0.5±0.2 | 0.4±0.2 | 0.2±0.1 |
| Moderate physical activity | 1.4±0.3 | 1.1±0.3 | 0.6±0.4 |
| Walking | 2.1±0.4 | 3.7±1.8 | 6.9±1.9[†] |
| Sitting | 45.3±5.7 | 56.9±5.5 | 50.4±7.7 |
| MET-minutes per week | 986±143 | 1162±377 | 1598±370 |

Data are means±SEM. [†]$p$<0.05 vs. lean. T2D, type 2 diabetes; IPAQ-SF, International Physical Activity Questionnaire Short Form.

**Table S2 - HIIT sessions durations, training intensity, workload and energy expenditure**

| | Lean | Obese | T2D |
|---|---|---|---|
| *n* | 16 | 15 | 13 |
| **Duration per session (min)** | | | |
| Week 1-4 | 28.5 | 28.5 | 28.5 |
| Week 5-8 | 54.5 | 54.5 | 54.5 |
| Week 1-8 | 41.5 | 41.5 | 41.5 |
| **Training intensity (%HR max)** | | | |
| Week 1-4: Peak intensity (peak %HRmax) | 96±0 | 96±0 | 96±0 |
| Week 5-8: Peak intensity (peak %HRmax) | 95±1 | 95±0* | 93±1**$^{\dagger\ddagger}$ |
| Week 1-8: Peak intensity (peak %HRmax) | 96±0 | 96±0 | 95±1 |
| Week 1-4: HIIT sessions incl. breaks | 80±1 | 82±1 | 82±1 |
| Week 5-8: HIIT sessions incl. breaks | 79±1** | 80±1* | 80±1** |
| Week 1-8: HIIT sessions incl. breaks | 80±1 | 81±1 | 81±1 |
| Week 1-4: Cycling intervals | 89±1 | 88±1 | 88±1 |
| Week 5-8: Cycling intervals | 88±1 | 87±1* | 85±1**$^{\dagger}$ |
| Week 1-8: Cycling intervals | 88±1 | 87±1 | 87±1 |
| Week 1-4: Rowing intervals | 88±1 | 88±1 | 87±1 |
| Week 5-8: Rowing intervals | 87±1* | 87±1 | 85±1* |
| Week 1-8: Rowing intervals | 88±1 | 87±1 | 86±1 |
| **Maximal cycling capacity (%)** | | | |
| Week 1-4 | 104±2 | 111±3$^{\dagger}$ | 112±2$^{\dagger}$ |
| Week 5-8 | 100±2* | 106±2*$^{\dagger}$ | 105±3* |
| Week 1-8 | 102±2 | 109±2$^{\dagger}$ | 108±2$^{\dagger}$ |
| **Workload (watt)** | | | |
| Week 1-4: Cycling intervals | 278±11 | 290±12 | 244±11$^{\dagger\ddagger}$ |
| Week 5-8: Cycling intervals | 286±11* | 297±11 | 252±11*$^{\dagger\ddagger}$ |
| Week 1-8: Cycling intervals | 282±11 | 294±12 | 248±11$^{\dagger\ddagger}$ |
| Week 1-4: Rowing intervals | 214±11 | 221±11 | 168±8$^{\dagger\ddagger}$ |
| Week 5-8: Rowing intervals | 229±11** | 244±13** | 185±9**$^{\dagger\ddagger}$ |
| Week 1-8: Rowing intervals | 221±11 | 230±12 | 176±8$^{\dagger\ddagger}$ |
| **Energy expenditure (kJ)** | | | |
| Week 1-4: Total during training intervals | 919±38 | 953±40 | 766±32$^{\ddagger}$ |
| Week 5-8: Total during training intervals | 1729±71** | 1759±91** | 1463±65**$^{\dagger\ddagger}$ |
| Week 1-8: Total during training intervals | 1311±54 | 1365±56 | 1100±46$^{\dagger\ddagger}$ |
| Week 1-4: During cycling intervals | 473±18 | 494±20 | 416±18 |
| Week 4-8: During cycling intervals | 873±32** | 909±34** | 770±33**$^{\dagger\ddagger}$ |
| Week 1-8: During cycling intervals | 669±25 | 698±103 | 589±25$^{\dagger\ddagger}$ |
| Week 1-4: During rowing intervals | 446±22 | 459±23 | 350±16$^{\dagger\ddagger}$ |
| Week 5-8: During rowing intervals | 855±42** | 910±47** | 693±35**$^{\dagger\ddagger\ddagger}$ |
| Week 1-8: During rowing intervals | 642±32 | 667±33 | 512±24$^{\dagger\ddagger}$ |

Data are means±SEM. **p*<0.05 and ***p*<0.001 week 5-8 vs. week 1-4, $^{\dagger}p<0.05$ and $^{\dagger\dagger}p<0.001$ vs. lean. $^{\ddagger}p<0.05$ and $^{\ddagger\ddagger}p<0.001$ vs. obese. T2D, type 2 diabetes; HRmax, heart rate max.
